# Reasoning Models for Text Mining in Oncology – a Comparison Between o1 Preview and GPT-4o

**DOI:** 10.1101/2024.12.06.24318592

**Authors:** Paul Windisch, Fabio Dennstädt, Christina Schröder, Daniel R. Zwahlen, Robert Förster

## Abstract

**Purpose:** Chain-of-thought prompting is a method to make a Large Language Model (LLM) generate intermediate reasoning steps when solving a complex problem to increase its performance. OpenAI’s o1 preview is an LLM that has been trained with reinforcement learning to create such a chain-of-thought internally, prior to giving a response and has been claimed to surpass various benchmarks requiring complex reasoning. The purpose of this study was to evaluate its performance for text mining in oncology.

**Methods:** Six hundred trials from high-impact medical journals were classified depending on whether they allowed for the inclusion of patients with localized and/or metastatic disease.

GPT–4o and o1 preview were instructed to do the same classification based on the publications’ abstracts.

**Results:** For predicting whether patients with localized disease were enrolled, GPT-4o and o1 preview achieved F1 scores of 0.80 (0.76 - 0.83) and 0.91 (0.89 - 0.94), respectively. For predicting whether patients with metastatic disease were enrolled, GPT-4o and o1 preview achieved F1 scores of 0.97 (0.95 - 0.98) and 0.99 (0.99 - 1.00), respectively.

**Conclusion:** o1 preview outperformed GPT-4o for extracting if people with localized and or metastatic disease were eligible for a trial from its abstract. o1 previews’s performance was close to human annotation but could still be improved when dealing with cancer screening and prevention trials as well as by adhering to the desired output format. While research on additional tasks is necessary, it is likely that reasoning models could become the new state of the art for text mining in oncology and various other tasks in medicine.

## Introduction

Ever since the development of the transformer architecture, Large Language Models (LLMs) have shown promising results for various text mining tasks in medicine, from extracting information from clinical notes to analyzing biomedical research publications.^1–3^ Most advances we have seen in recent years have been based on a “bigger is better approach” that relies on increasing the learning data and parameter count resulting in increasingly large models.^4^

A method that has been employed by users and developers of LLMs alike is chain-of-thought prompting, i.e. making a model generate intermediate reasoning steps when solving a complex problem to increase its performance at inference without having to resort to a bigger model.^5^ With the release of its o1 preview model, OpenAI has for the first time published a model that has been trained with reinforcement learning to create such a chain-of-thought internally, prior to giving a response. The company claims that the model surpassed various previously established benchmarks related to coding, math, and answering science questions, not just those set previously by OpenAI models but by LLMs in general.^6^

Doing text mining in oncology is a task that frequently requires complex reasoning. In a previous publication, our group assessed the performance of various models on processing abstracts from oncological randomized clinical trial (RCT) publications and extracting whether each trial allowed for patients with localized and/or metastatic disease to be included.^7^ This task is not straightforward as the information can be conveyed in various ways, such as the UICC or AJCC TNM stage (which varies between tumor entities), or ambiguous keywords such as “advanced” or “extensive”.^8^

The goal of this project was therefore to evaluate the performance of OpenAI’s o1 preview against its flagship model GPT-4o which had previously achieved performance that was similar to dedicated models with task-specific pretraining.^7^ Our hypothesis was that o1 preview would be able to surpass GPT-4o’s performance, driven by an improvement on abstracts that require more complex reasoning to make a prediction.

## Methods

We used a previously annotated dataset that contained 600 randomly chosen publications from seven major journals (British Medical Journal, JAMA, JAMA Oncology, Journal of Clinical Oncology, Lancet, Lancet Oncology, New England Journal of Medicine) published between 2005 and 2023.^9^ Trials were labeled depending on whether they enrolled patients with localized disease, metastatic disease, neither (in case of cancer prevention and screening trials), or both. Annotation was performed by a single author (P.W.) using the tool prodigy (v. 1.13.1). Since using GPT-4o and o1 preview does not require task-specific pre-training, the full dataset was used to evaluate the performance of the models.

Calls to the models were made via an application programming interface (API). For GPT-4o (OpenAI, San Francisco, United States), the system prompt was the following: “You will be provided with the abstract of a cancer clinical trial. Your task will be to classify, if patients with metastatic disease were eligible for the trial or not. In addition, you will be asked to classify, if patients with localized or locally advanced disease, i.e. cancer that has not formed distant metastases, were eligible for the trial. Your response should be a list of two boolean values (True or False), the first indicating if patients with metastatic disease were eligible and the second indicating if patients with localized or locally advanced disease were eligible. The list should be enclosed in brackets and separated by a comma, e.g. [True, False].”

The user prompt was the respective abstract. As o1 preview does not support a system prompt at the time of writing, both the instructions and the respective abstract were sent as the user prompt.

The respective snapshots that were used were gpt-4o-2024-08-06 for GPT-4o and o1-preview-2024-09-12 for o1 preview.

The temperature was set to 1.0 to allow for a direct comparison as this is the only temperature setting that is supported by o1 preview at the time of writing. No max_tokens or max_completion_tokens limit was used to avoid restricting o1’s reasoning capabilities as tokens generated during the chain-of-thought process count towards the max_completion_tokens limit.

After obtaining the responses from the LLMs, answers were formatted using regular expressions and performance metrics (accuracy, precision, recall, and F1 score) were calculated. Due to the relatively high number of false negative predictions by both models when predicting if people with localized disease were enrolled in a trial, we conducted a manual inspection of these trials and quantified the presence of certain keywords.

The cost for inference was assessed by using OpenAI’s usage dashboard.

All analyses were performed in python (version 3.11.5) using, among others, the pandas (version 2.1.0), and openai (version 1.40.3) packages. The code and data including the raw LLM responses are available from github (https://github.com/windisch-paul/o1) and Dryad (reviewer link: http://datadryad.org/stash/share/2xRq2jz0e8dbp_EU2KsZVeLFARb-j0C0YZI60wBD7qk).

## Results

The distribution of trials that allowed for the inclusion of patients with localized and/or metastatic disease is presented in Figure 1a. While 73.7% of trials enrolled patients with localized disease, 59.2% of trials allowed patients with metastatic disease. Screening or prevention trials that did not enroll patients with an active cancer accounted for 5% of trials in the dataset.

**Figure 1.**
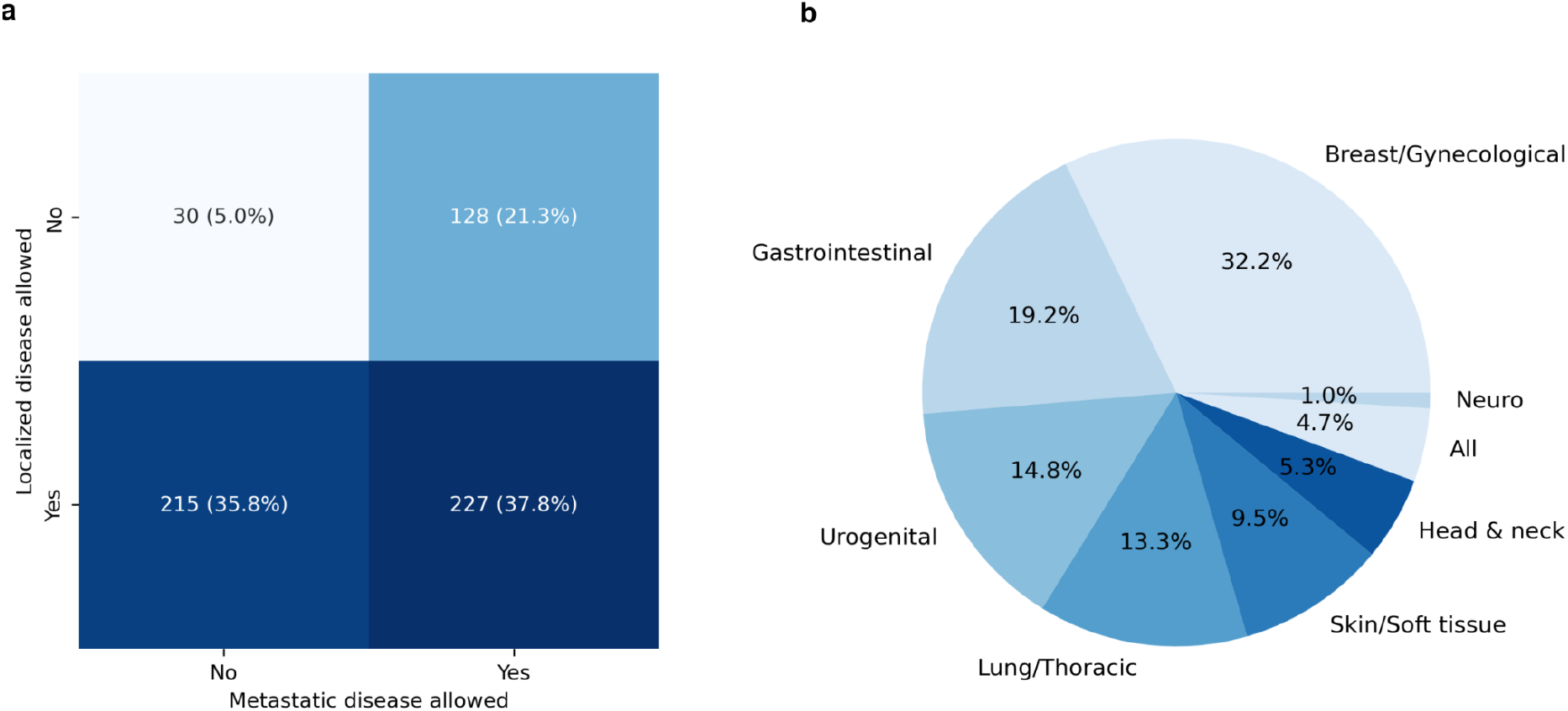
**a)** Distribution of trials that allowed for the inclusion of patients with localized and metastatic disease in the dataset **b)** Distribution of grouped tumor entities in the dataset. Trials that allowed for the inclusion of patients with any tumor entity e.g. trials for patients with painful bone metastases from any primary site are grouped under “All”.

The distribution of grouped tumor entities is presented in Figure 1b. Breast and gynecological tumors were investigated in 32.2% of trials, followed by gastrointestinal tumors in 19.2%, urogenital tumors in 14.8% and lung/thoracic tumors in 13.3% of trials.

The total cost for making predictions for all trials in the dataset with GPT-4o was USD 1.23 and USD 44.01 for o1, which is equivalent to a cost per abstract of 0.21 cent per trial for GPT-4o and 7.3 cent for o1 preview.

GPT-4o returned the desired output format (i.e. two boolean values enclosed in brackets separated by a comma) for every trial. For o1 preview, the desired output format was returned for 86% (n = 516) of trials. For the remaining 14% (n = 84) of trials the output was more verbose, i.e. some of the chain-of-thought was present in the response. However, in all but 1.8% (n = 11) of trials, the desired output format was also present in and could be extracted from the verbose output.

For predicting whether patients with localized disease were enrolled, GPT-4o and o1 preview achieved F1 scores of 0.80 (0.76 - 0.83) and 0.91 (0.89 - 0.94), respectively. For predicting whether patients with metastatic disease were enrolled, GPT-4o and o1 preview achieved F1 scores of 0.97 (0.95 - 0.98) and 0.99 (0.99 - 1.00), respectively. The full performance metrics are presented in Table 1. The confusion matrices are presented in Figure 2. The performance per tumor entity group is presented in Table 2. The largest discrepancies in F1 scores for predicting whether people with localized disease were enrolled were observed for skin, lung, and breast tumors respectively (F1 score differences: 0.23, 0.20, and 0.11). For the false negative predictions made by o1, 60% (n = 39) of abstracts contained the word “advanced” while no abstract contained the phrase “locally advanced”. 26.2% (n = 17) of abstracts contained the word “recurrent”.

**Table 1.**
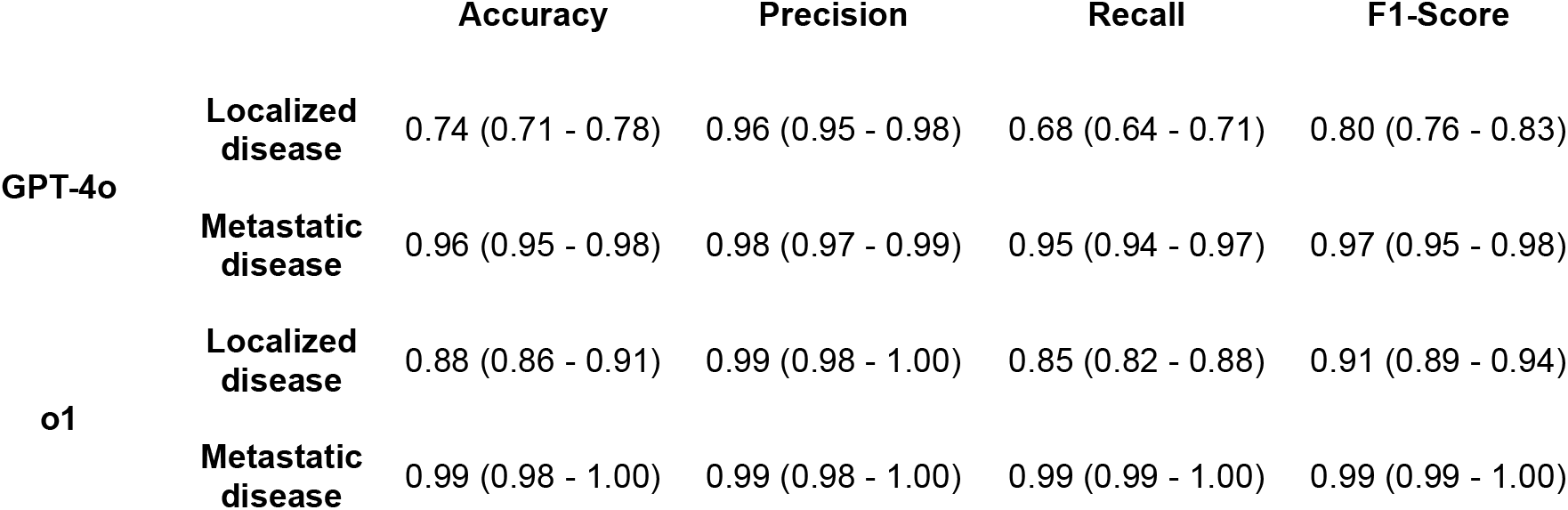
Performance metrics. Numbers in parentheses indicate the 95% confidence intervals; *ML = Machine Learning*.

|  |  | Accuracy | Precision | Recall | F1-Score |
| --- | --- | --- | --- | --- | --- |
| GPT-4o | Localized disease | 0.74 (0.71 - 0.78) | 0.96 (0.95 - 0.98) | 0.68 (0.64 - 0.71) | 0.80 (0.76 - 0.83) |
|  | Metastatic disease | 0.96 (0.95 - 0.98) | 0.98 (0.97 - 0.99) | 0.95 (0.94 - 0.97) | 0.97 (0.95 - 0.98) |
| o1 | Localized disease | 0.88 (0.86 - 0.91) | 0.99 (0.98 - 1.00) | 0.85 (0.82 - 0.88) | 0.91 (0.89 - 0.94) |
|  | Metastatic disease | 0.99 (0.98 - 1.00) | 0.99 (0.98 - 1.00) | 0.99 (0.99 - 1.00) | 0.99 (0.99 - 1.00) |

**Table 2.** F1 scores of the respective models for the largest tumor entity groups.

|  |  | Breast/<br>Gynecological | Gastro-<br>intestinal | Urogenital | Lung/<br>Thoracic | Skin/<br>Soft tissue | Head/<br>Neck |
| --- | --- | --- | --- | --- | --- | --- | --- |
| GPT-4o | Localized<br>disease | 0.84 | 0.78 | 0.89 | 0.59 | 0.71 | 0.90 |
|  | Metastatic<br>disease | 0.94 | 1.00 | 0.99 | 0.99 | 0.96 | 1.00 |
| o1 | Localized<br>disease | 0.95 | 0.85 | 0.95 | 0.79 | 0.94 | 0.90 |
|  | Metastatic<br>disease | 0.98 | 0.98 | 0.99 | 0.99 | 1.00 | 0.92 |

**Figure 2.**
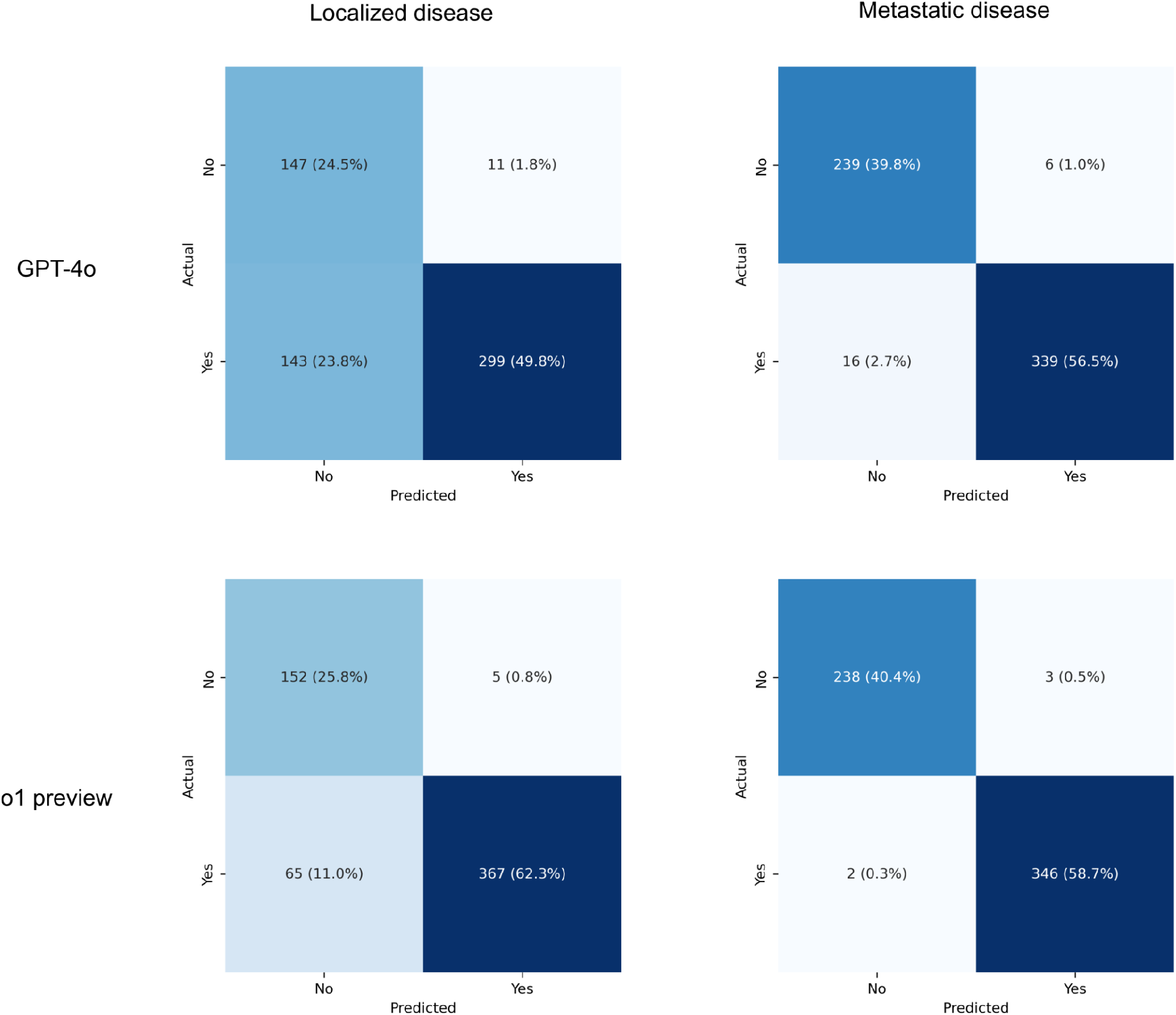
Confusion matrices for the performance of GPT-4o (top) and o1 (bottom) on predicting whether patients with localized disease (left) and/or metastatic disease (right) were eligible for a trial.

## Discussion

On our dataset of oncology RCTs, o1 preview surpassed GPT-4o’s performance when predicting whether patients with localized and/or metastatic disease were eligible based on the publication’s abstract. While the performance was more similar and almost on par with the human annotator for predicting if patients with metastatic disease were eligible (F1 scores of 0.97 and 0.99 for GPT-4o and o1 preview, respectively), the difference was more pronounced for localized disease (F1 scores of 0.80 and 0.91 for GPT-4o and o1 preview, respectively).

Possible explanations for this become apparent when reviewing incorrect classifications. The few misclassified examples regarding metastatic disease, especially those also misclassified by o1 preview, mostly did not provide information regarding whether patients with metastatic disease were enrolled in the trial. Sometimes this information was mentioned in the title of the publication, but the LLM was not provided with the title when making a prediction.

Misclassifications for localized disease were much more frequent and mostly related to ambiguous language. This is indicated by the high proportion of abstracts misclassified by o1 preview that contained the words “advanced” (60% of false negatives, n = 39) or “recurrent” (26.2% of false negatives, n = 17). Advanced might mean metastatic but might also mean locally advanced. Recurrent could refer to a local or a distant recurrence. These phrases frequently caused the human annotator to read the full publication to annotate the ground truth which the LLM could not do. Therefore, the performance of o1 preview might actually be even closer to human performance considering the different information available to human vs. AI for the classification. The improved performance compared to GPT-4o could be an indication that o1 preview was more able to handle this ambiguity, potentially by using other information in the text to make its prediction. As an example, the endpoints of a study could also have been used to assess which patients were allowed in a trial, such as disease free or metastasis free survival being more frequently used in trials of localized disease.^10^

These more subtle clues require the kind of more complex reasoning that o1 preview is supposed to employ.

A concept that even o1 preview occasionally struggled with were screening and prevention trials which accounted for all false positives when predicting if patients with localized disease were eligible (0.8%, n = 5). In addition, the fact that no prediction in the correct format was made for 1.8% (n = 11) of abstracts and that the output was more verbose than requested in 14% (n = 84) of abstracts leaves additional room for improvement. Whether the substantially higher price point warrants the improved performance compared to GPT-4o depends on the respective task the model is used for.

While our study represents, to the best of our knowledge, the first evaluation of o1 preview for a text mining task in medicine, our findings are in line with other studies of its capabilities.

Zhong and colleagues compared o1 preview to a variety of other OpenAI models for various tasks and found an improved performance with o1 preview for generating radiology reports.^11^ Chang and colleagues as well as Goto and colleagues found an improved performance with o1 preview compared to GPT-4o for answering multiple choice questions on the Taiwan psychiatry licensing examination and the Japanese ‘Operations Chief of Radiography With X-rays’ certification, respectively.^12, 13^ Nori, Usuyama et al. also found an improved performance for various medical multiple choice question benchmarks including the United States Medical Licensing (USMLE) Sample Exam and Self Assessment while highlighting that o1 preview even outperformed GPT-4o when the latter was used with sophisticated prompting techniques and the former was not.^14^ A slightly higher rate of hallucinations compared to GPT-4o (21.3% vs. 20.0%) was reported by Erdem and colleagues who investigated these models for tasks associated with financial literature which is in line with the increased deviations from the desired output format in our study.^15^

A limitation of this study is that it evaluated the performance on abstracts from seven journals that all use structured abstracts. However, due to the breadth of data that these models have encountered during training, the likelihood of the findings also applying to other journals and unstructured abstracts seems high. In addition, since most of the studies had been published prior to the LLMs being trained, there is at least the theoretical chance that the models have been trained on both the abstract and the associated publication so that they could make their prediction not only based on the abstracts that they were provided but also by recalling details of the publication. However, this risk is mitigated by the fact that at least some of the journals lock their full publications behind paywalls.

The strengths of this study include a fairly large and heterogenous test set as well. Furthermore, all data and code are publicly available.

In conclusion, o1 preview outperformed GPT-4o for extracting if people with localized and or metastatic disease were eligible for a trial from its abstract. o1 preview’s performance was close to human annotation but could still be improved when dealing with cancer screening and prevention trials as well as by adhering to the desired output format. While research on additional tasks is necessary, it is likely that reasoning models could become the new state of the art for text mining in oncology and various other tasks in medicine.

## Data Availability

All data and code used to obtain this study's results have been uploaded to https://github.com/windisch-paul/o1. The data has also been uploaded to Dryad but is private while the paper is undergoing review.

